# Ischemic cerebral lesions after Carotid Stenting versus Carotid Endarterectomy: A Systematic review and Meta-Analysis

**DOI:** 10.1101/2024.03.18.24304513

**Authors:** Georgios Loufopoulos, Vasiliki Manaki, Panagiotis Tasoudis, Andreas Stylianos Meintanopoulos, George Kouvelos, George Ntaios, Konstantinos Spanos

## Abstract

**Background:** Recent randomized controlled trials have demonstrated similar outcomes in terms of ischemic stroke incidence after carotid endarterectomy (CEA) or carotid artery stenting (CAS) in asymptomatic carotid disease, while CEA seems to be the first option for symptomatic carotid disease. The aim of this meta-analysis is to assess incidence of silent cerebral microembolization detected by Magnetic Resonance Imaging (MRI) following these procedures.

**Methods:** A systematic search was conducted using PubMed, Scopus and Cochrane databases including comparative studies involving symptomatic or asymptomatic patients undergoing either CEA or CAS, and reporting on new cerebral ischemic lesions in post-operative MRI. The primary outcome was the newly detected cerebral ischemic lesions. Pooled effect estimates for all outcomes were calculated using the random-effects model. Pre-specified random effects meta-regression and subgroup analysis were conducted to examine the impact of moderator variables on the presence of new cerebral ischemic lesions.

**Results:** 25 studies reporting on total 1827 CEA and 1500 CAS interventions fulfilled the eligibility criteria. The incidence of new cerebral ischemic lesions was significantly lower after CEA comparing to CAS, regardless of the time of MRI assessment (first 24 hours; OR: 0.33, 95% CI: 0.17-0.64, p<0.001), (the first 72 hours, OR: 0.25, 95% CI 0.18-0.36, p<0.001), (generally within a week after the operation; OR: 0.24, 95% CI: 0.17-0.34, p<0.001). Also, the rate of stroke (OR: 0.38, 95% CI: 0.23-0.63, p<0.001) and the presence of contralateral new cerebral ischemic lesions (OR: 0.16, 95% CI 0.08-0.32, p<0.001) were less frequent after CEA. Subgroup analysis based on the study design and the use of embolic protection device during CAS showed consistently lower rates of new lesions after CEA.

**Conclusions:** CEA demonstrates significant lower rates of new silent cerebral microembolization, as detected by MRI in postoperative period, compared to CAS.

## Introduction

Both carotid endarterectomy (CEA) and carotid artery stenting (CAS) reduce the risk of stroke or transient ischemic attack (TIA) with relatively low morbidity and mortality in people with specific comorbidities and features, as well as suitable carotid lesions^1–4^. One of the main risks during these procedures is embolization. CEA is the preferred treatment for both symptomatic and asymptomatic individuals; nonetheless, in high-risk patients CAS is an alternative therapeutic option^5^. Despite its less invasive nature, CAS seems to have higher risk of periprocedural embolization compared to CEA most likely due to manipulation of endovascular devices in the aortic arch or in friable plaques^6–8^.

While stroke/TIA after carotid revascularization is comparatively rare, subclinical microembolization was found to occur more frequently^9–11^. Silent microembolization episodes may nowadays be recognized more easily than in the past, with transcranial Doppler and diffusion weighted Magnetic Resonance Imaging (DWI-MRI) providing the highest diagnostic accuracy^5,12^. DWI-MRI is a high-sensitivity imaging tool capable of detecting silent infarcts following carotid revascularization and providing important information on the safety of various procedures.

Various studies reported the occurrence of confirmed DWI-MRI silent microembolization following CEA or CAS^13–15^. To synthesize the most recent data from the current literature on the incidence of silent cerebral microembolization following CEA and CAS, we carried out a systematic review and meta-analysis of studies that used DWI-MRI for microembolization detection before and after carotid revascularization. Our research attempts through rigorous mathematical methodology to provide concrete evidence and compare the most updated incidence of silent post-operative microembolization between CEA and CAS.

## Material and Methods

### Study design and inclusion/exclusion criteria

This systematic review and meta-analysis were performed according to the PRISMA (Preferred Reporting Items for Systematic Reviews and Meta-analyses) guidelines^16^and were prospectively registered in the International Prospective Register of Systematic Review (PROSPERO) database (registration number: CRD42023494604). We applied the PICO (Population/Participants, Intervention, Comparison, and Outcome) criteria to define our research question:

1. Population/Participants: Adult patients, symptomatic or asymptomatic, with carotid stenosis undergoing operation and performing pre and post-procedural MRI.
2. Intervention: CEA
3. Comparison group: CAS
4. Outcomes: The primary assessed outcome was the new cerebral ischemic lesions shown in the post-procedural MRI. The secondary outcomes were the presence of contralateral cerebral ischemic lesions and stroke.

We also performed subgroup analyses according to the time of postoperative MRI, the study design, and the use of embolic protection device with the CAS technique. Pre-specified random effects meta-regression analysis was conducted to examine the impact of pre-operative symptoms, hypertension, smoking status and diabetes on the presence of new cerebral ischemic lesions. Stroke was defined as either ipsilateral or contralateral, cortical or vertebrobasilar, ischemic or hemorrhagic strokes, during the first 30 postoperative days.

Original clinical studies, including both randomized trials and non-randomized prospective/retrospective comparative studies, reporting on the outcomes of interest in patients undergoing CEA and CAS, were deemed eligible for inclusion. The exclusion criteria for the present systematic review were defined as follows: (i) irrelevant articles, (ii) non-comparative studies (<2 study arms), (iii) studies not directly comparing CEA and CAS for the outcomes of interest, (iv) animal and in-vitro studies, (v) case reports, (vi) narrative or systematic reviews and meta-analyses, letters to the editor and comments, (vii) non-English language articles and (viii) published abstracts with no published full text. Ultimately the studies identified were assessed for overlap. In cases where multiple studies reported on the same population, only the larger study or the one with the best quality of data was included in the present meta-analysis.

### Literature search strategy

Eligible studies were identified by searching through the MEDLINE (via PubMed), Scopus, and Cochrane Library databases (last search: November ^29th^, 2023) using the algorithm: (CEA OR (endarterectomy, carotid[MeSH Terms])) OR endarterectom*) AND ((stent[MeSH Terms]) OR CAS OR ("carotid stent") OR ("carotid stenting") OR (angiopl*)) AND ("diffusion weighted imaging" OR DWI OR MRI OR ("magnetic resonance imaging") OR ("ischemic lesion") OR (diffusion magnetic resonance imaging[MeSH Terms])) AND ((arterial disease, carotid[MeSH Terms]) OR (carotid arteries[MeSH Terms]) OR (caroti*) OR (carotid stenosis[MeSH Terms]) OR (carotid stenoses) OR (arterial disease, carotid[MeSH Terms]) OR "carotid surgery"). No search filters were applied to our search. Title and abstract screening and full-text eligibility were assessed by two independent investigators. Any disagreement was resolved after a discussion with a third reviewer. We also searched the reference list of the included studies for potentially eligible studies using the snowball methodology^17^. The Covidence reference and article manager software was used for all stages of the database search and study selection^18^.

### Data extraction and assessment of the risk of bias

Two investigators independently extracted the data into a pre-designed standardized form. Patients’ baseline characteristics as well as peri-operative data and post-operative outcomes of interest were collected. All the outcomes of interest occurred in the first 30 days after the operation.

The Risk of Bias in Non-Randomized Studies of Interventions tool (ROBINS-I) and the revised Cochrane risk of bias tool for randomized trials (RoB 2) were systematically used to assess included studies for risk of bias in non-randomized studies and in RCTs, respectively^19,20^. The papers and their characteristics were classified into low, moderate, serious, or critical risk of bias with ROBINS-I tool and low, some concerns or high risk of bias with RoB2 tool. Two independent reviewers assessed the risk for bias. When there was disagreement, a third reviewer checked the data and made the final decision.

### Statistical Analysis

#### Data Pooling

Continuous variables were summarized using means and standard deviations (SDs), while categorical variables were summarized by frequencies and percentages. The Hozo et al. and the Wan et al. methods were used to estimate the means and standard deviations of continuous variables whenever medians and ranges^21^ and median and interquartile ranges were provided^22^, respectively. Data were extracted and entered into tables and the outcomes were analyzed cumulatively.

#### Meta-Analysis

We calculated odds ratios (ORs) and 95% confidence intervals (CIs) for all individual studies based on the extracted data, using 2 x 2 tables for each categorical outcome. OR >1 indicated that the outcome was more frequently present in the CEA group. A treatment arm continuity correction was adopted in studies with zero cell frequencies^23^. Between-study heterogeneity was assessed through Cochran Q statistic and by estimating I^2^. I^2^ greater than 50% and *p<*0.1 indicated significant heterogeneity. Due to the significant between-study clinical heterogeneity, we used the random-effects model (DerSimonian–Laird) to calculate the pooled effect estimates for all outcomes^24^. A forest plot for each outcome was used to display the pooled estimates graphically. Publication bias was assessed via funnel plots. Egger’s test was used when at least 10 studies were included in the analysis of each outcome of interest and p < 0.10 was considered statistically significant, indicating possible publication bias. Subgroup analysis was performed according to the time of postoperative MRI, the study design and the use of embolic protection device with the CAS technique. Pre-specified random effects meta-regression analysis was conducted to examine the impact of moderator variables on the presence of new cerebral ischemic lesions. Specifically, using this technique we attempted to assess the effect of pre-operative symptomatic status, hypertension, smoking and diabetes on the incidence of newly detected lesions. These moderator variables were expressed as difference in rate of occurrence in the CEA versus the CAS group. Statistical analysis was performed using Stata/SE version 18 (Stata Corp, College Station, TX, USA).

## Results

### Study and Patient characteristics

The literature search yielded 804 potentially eligible articles after duplicates were removed, of which 66 underwent full-text evaluation. A total of 21 studies were excluded due to overlapping populations. Eight analyses of the International Carotid Stenting study (ICSS) were identified^25–32^. Among these, we selected the one with the largest number of patients and reporting granular data of the outcomes of interest^27^. We used the same methodology to conclude to the 1 study^33^ and exclude the other 6 analyses reporting on patients from the Veterans Affairs Palo Alto Health Care System^34–39^. In 1 study we preferred to use in our analysis the data from the propensity matched pair of patients, and not from the entire original population^40^. A non-English written study was also excluded from our pooled analysis because in the Spanish language the critical assessment of the study’s quality may be inaccurate^41^. Finally, 25 studies reporting on a total of 3327 patients undergoing CEA and CAS for carotid stenosis, fulfilled the inclusion criteria and were included in our quantitative data analysis, as summarized in the PRISMA flowchart **(Figure 1)**^27,33,40,42–63^. Among them, a total of 3366 procedures were reported, 1861 CEA, and 1505 CAS. Pre and post-operative MRI was performed in total 1827 of 1861 CEA procedures and 1500 of 1505 CAS interventions. The baseline characteristics of the included studies, the patient’s demographics, and the perioperative outcomes are summarized in **Table 1**, **Supplementary Table 1** and **Supplementary Table 2**, respectively.

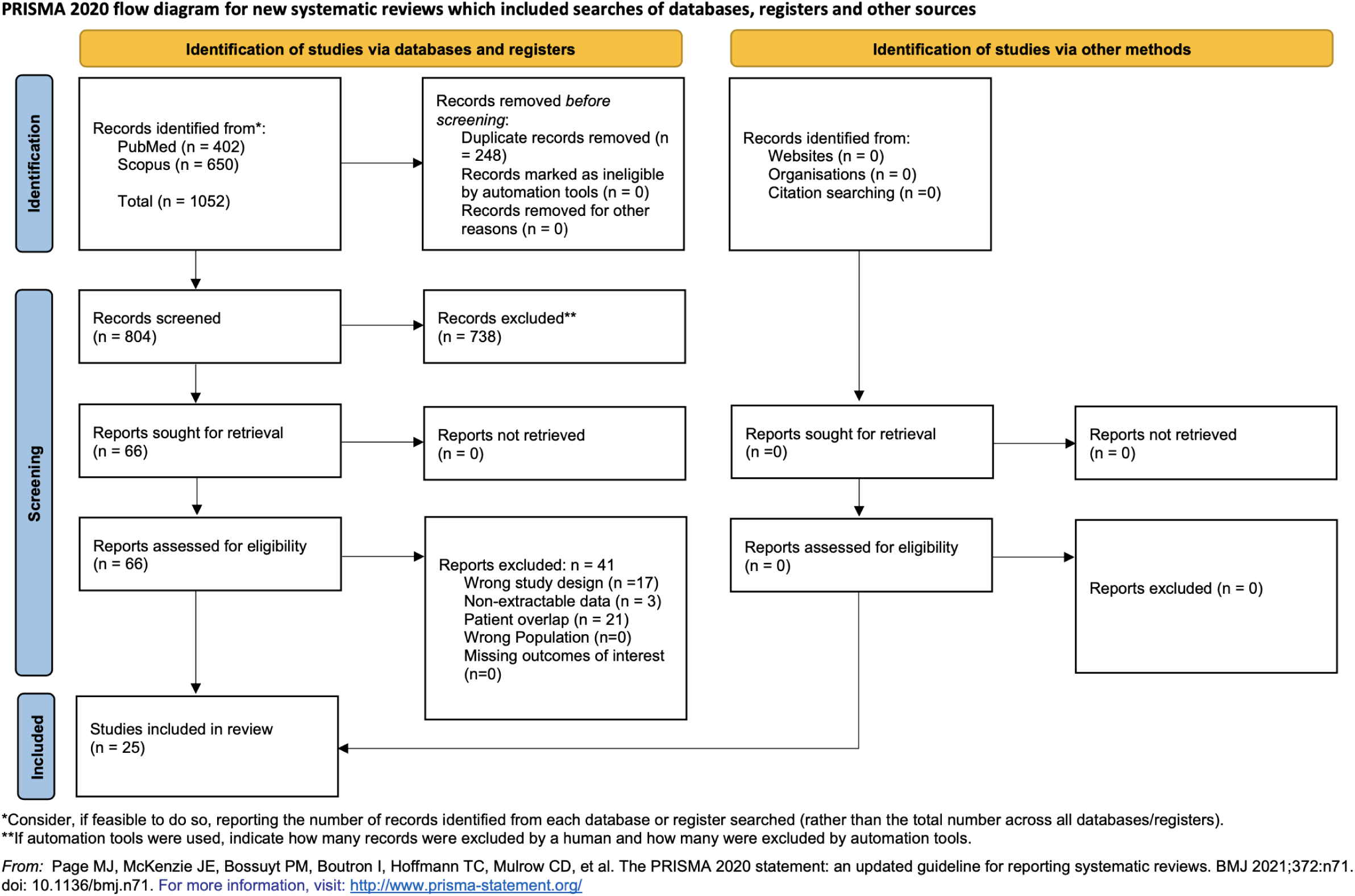

**Table 1.** Study characteristics.

| Author | Year | Journal | Center | Country | Design | Study period | Total | CEA | CAS |
| --- | --- | --- | --- | --- | --- | --- | --- | --- | --- |
| Lacroix V et al. | 2007 | European Journal of Vascular and Endovascular Surgery | Cliniques Universitaires Saint-Luc, Service de Chirurgie cardiovasculaire et thoracique | Belgium | Prospective cohort | - | 121 | 60 | 61 |
| Yan D et al. | 2018 | Annals of Vascular Surgery | Shanghai Ninth People's Hospital and Zhongshan Hospital, Shanghai | China | Retrospective | January 2012-June 2017 | 92 | 38 | 54 |
| Kuliha M et al. | 2015 | British Journal of Surgery | University Hospital Ostrava, Ostrava | Czech Republic | Randomized Control Trial (RCT) | October 2010-July 2014 | 150 | 77 | 73 |
| Poppert H et al. | 2004 | Journal of Neurology | Klinikum Rechts der Isar Muenchen | Germany | Prospective cohort | - | 129 | 88 | 41 |
| Wasser K et al. | 2011 | Journal of Vascular Surgery | University of Göttingen, Göttingen | Germany | Prospective cohort | - | 55* | 31 | 24 |
| Mihaly Z et al. | 2023 | Journal of Cardiovascular Development and Disease | Department of Vascular and Endovascular Surgery, Heart and Vascular Center, Semmelweis University, Budapest | Hungary | Prospective cohort | 1 January 2019-31 January 2021 | 42 | 21 | 21 |
| Faraglia V. et al. | 2007 | The Journal of Cardiovascular Surgery | Sant Andrea Hospital La Sapienza University of Rome Seconde Medical school, Rome | Italy | Prospective cohort | December 2005-January 2007 | 75 | 40 | 35 |
| Felli MM et al. | 2012 | International Angiology | Paride Stefanini Vascular surgery Polilinico Umberto 1, Rome | Italy | Prospective cohort | January 2008-January 2011 | 300 | 150 | 150 |
| Gabrielli R et al. | 2023 | Annals of Vascular Surgery | Unit of Vascular, Endovascular and Emergency Vascular Surgery, "S. Eugenio" Hospital, Rome | Italy | Prospective cohort | February 2018-June 2021 | 211 | 116 | 95 |
| Akutsu N. et al. | 2012 | American Journal of Neuroradiology | Department of Neurosurgery, Kobe University Graduate School of Medicine, Kobe | Japan | Prospective cohort | August 2006-February 2011 | 104 | 63 | 41 |
| Fukumitsu R et al. | 2017 | World Neurosurgery | Department of Neurosurgery, Kurashiki Central Hospital, Okayama | Japan | Retrospective | 2004-2013 | 295 | 181 | 114 |
| Iihara K et al. | 2006 | Journal of Neurosurgery | National Cardiovascular Center, Osaka | Japan | Retrospective | September 1998 and August 2004 | 231 | 139 | 92 |
| Mitsuoka H et al. | 2011 | Annals of Vascular Surgery | Shizuoka Red Cross Hospital, Shizuoka, Shizuoka | Japan | Prospective cohort | July 2007-May 2010 | 45 | 25 | 20 |
| Miyawaki S et al. | 2014 | Neurologia Medico Chirurgica | Aizu Chuo Hospital, Aizuwakamatsu, Fukushima | Japan | Retrospective | January 2000-September 2010 | 320 | 259 | 61 |
| Okamoto T et al. | 2022 | Surgical Neurology International | Kyoto Prefectural University of Medicine Graduate School of Medical Science, Kyoto | Japan | Retrospective | January 2012-May 2020 | 140 | 38 | 102 |
| Yamada K et al. | 2011 | Atherosclerosis | Gifu University Graduate School of Medicine, Gifu | Japan | Retrospective | June 2007-April 2010 | 81 | 25 | 56 |
| Roh H et al. | 2005 | American Journal of Neuroradiology | Samsung Medical Center, Sungkyunkwan University School of Medicine, Seoul | Korea | Prospective cohort | July 1998-February 2001 | 48 | 26 | 22 |
| Cho H et al. | 2014 | Stroke | san Medical Center, University of Ulsan College of Medicine, Seoul | South Korea | Prospective cohort | July 2007-December 2009 | 45 | 29 | 16 |
| Flach H et al. | 2004 | Journal of Endovascular Therapy | Vascular Surgery, Erasmus Medical center , Rotterdam | Netherlands | Prospective cohort | February 2001- November 2002 | 44 | 23 | 21 |
| Skjelland M et al. | 2009 | Stroke | Rikshospitalet University Hospital, Oslo | Norway | Prospective cohort | - | 91** | 61 | 30 |
| Latacz P et al. | 2019 | Polish archives of internal medicine | University Hospital, Jagiellonian University Medical College, Kraków | Poland | Randomized Control Trial (RCT) | May 2015-March 2018 | 31 | 14 | 17 |
| Bonati L et al. | 2010 | The Lancet Neurology | ICSS-MRI substudy, Amsterdam, Basel, Newcastle, Rotterdam, Sheffield, Utrecht | Switzerland, UK, Netherlands | Randomized Control Trial (RCT) | May 2001-October 2008 | 231 | 107 | 124 |
| Posacioglu H et al. | 2008 | Texas Heart Institute Journal | Ege University Hospital Bornova-Izmir | Turkey | Prospective cohort | February 2003-March 2005 | 115 | 59 | 56 |
| Sabat J et al. | 2018 | Journal of Vascular Surgery | University of Arizona, 1501 N Campbell Ave, Tucson | USA | Prospective cohort | July 2011-October 2016 | 202 | 95 | 107 |
| Zhou W et al. | 2009 | Journal of Vascular Surgery | Veterans Affairs Palo Alto Health Care System | USA | Retrospective | July 2004-December 2008 | 168 | 100 | 68 |

**Abbreviations:** CEA: Carotid endarterectomy; CAS: Carotid Artery Stenting
\*MRI was available in 49 procedures, 28 CEA and 21 CAS
\*\*MRI was performed in 58 procedures, 30 CEA and 28 CAS

### Study quality and publication bias assessment

Of the 25 included studies in the final analysis, 22 were non-randomized studies^33,40,43–56,58–63^ (7 retrospective and 15 prospective) and 3 were randomized controlled trials^27,42,57^. In 1 of the 22 non-randomized studies, the propensity score matching technique was used to adjust preoperative patients’ demographic and comorbidity data^40^. Seven non-randomized studies were considered with serious risk of bias: 6 due to confounding and 1 due to missing data **(Supplemental Figure 1)**. The RCTs were deemed of some concerns in terms of randomization, except one that was detected with high risk of bias **(Supplemental Figure 2)**. The funnel plots assessing for publication bias for the primary outcome of the presence of new ischemic cerebral lesions and the stroke rate are presented in **Supplemental Data File 1**. No publication was identified with the Egger’s test and through visual assessment of the funnel plots.

### Outcomes of interest

#### New cerebral ischemic lesions in post-operative MRI

##### 24-hours post-procedure

In 8 studies the MRI was performed during the first 24 hours ^44–46,49,50,54,56,57^. The rate of new cerebral ischemic lesions was significantly lower in the CEA group (14.3%, 112/782) compared to the CAS group (41.3%, 204/494) (OR: 0.33, 95% CI: 0.17-0.64, p<0.001). The statistical heterogeneity was considerable (I^2^=78.75%) **(Figure 2)**. The presence of contralateral new ischemic cerebral lesions was reported in 5 studies^44,46,49,56,57^. The incidence of contralateral ischemic lesions was significantly lower in the CEA group (0.5%, 2/399) compared to CAS (8.7%, 27/310) (OR: 0.13, 95% CI 0.04-0.42, p<0.001). The statistical heterogeneity was low (I^2^ = 0.00%) **(Supplemental Figure 7)**.

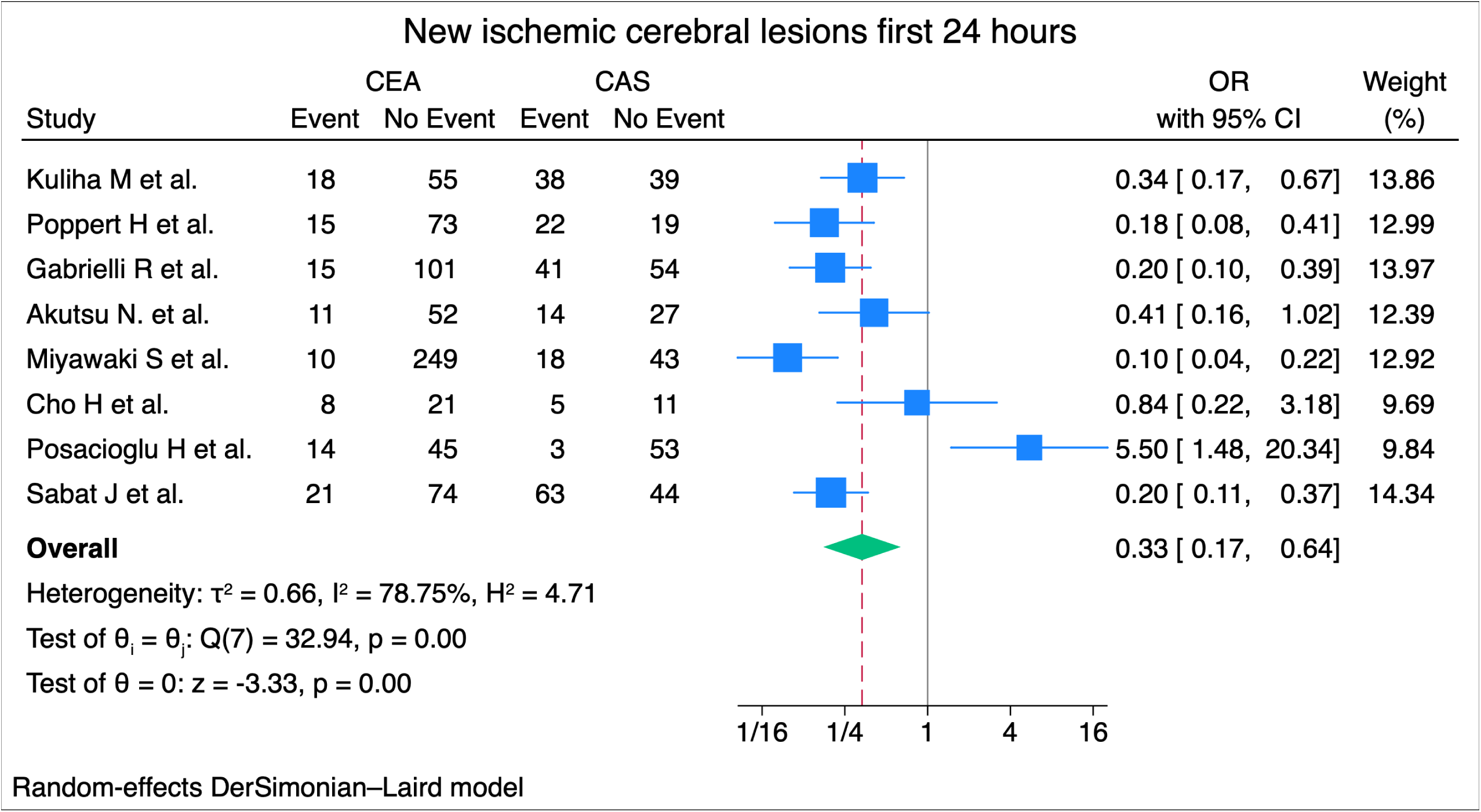

##### 72-hours post-procedure

The MRI was performed within 72 hours after the procedure in 22 studies^27,33,40,42–51,54–57,59–63^. The incidence of new cerebral ischemic lesions was significantly lower after the CEA procedures (13.6%, 237/1738) compared to CAS (40.4%, 567/1404) (OR: 0.25, 95% CI 0.18-0.36, p<0.001). The statistical heterogeneity was substantial (I^2^= 68.74%) **(Figure 3)**.

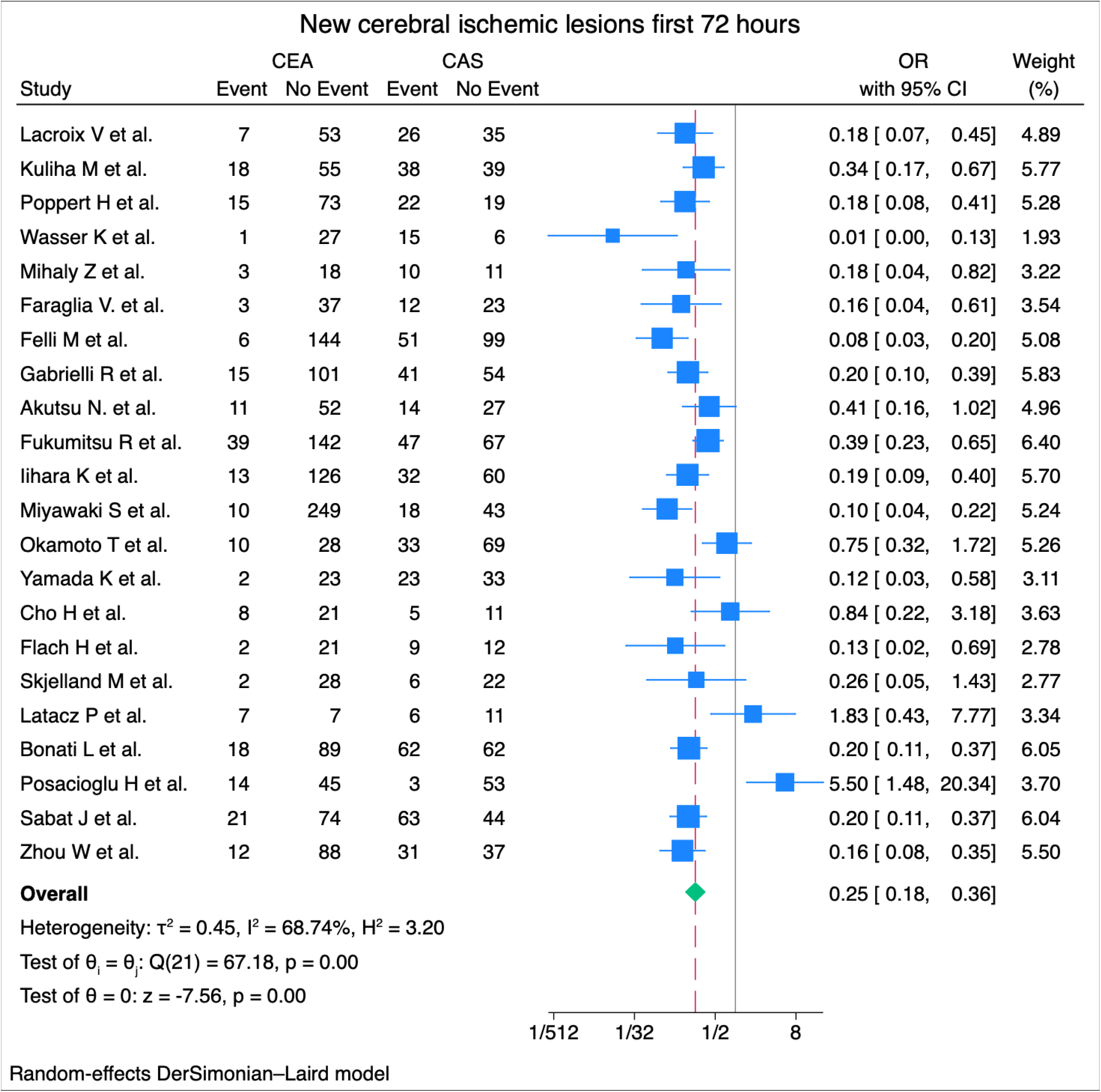

##### One week post-procedure

All 25 studies performed the MRI within a week after the operation ^27,33,40,42–63^. The rate of new cerebral ischemic lesions was significantly lower in the CEA group (13.7%, 251/1827) compared to the CAS group (41.1%, 616/1500) (OR: 0.24, 95% CI: 0.17-0.34, p<0.001). The statistical heterogeneity was substantial (I^2^= 66.85%) **(Figure 4)**. Eleven studies reported data regarding the presence of contralateral ischemic cerebral lesions^27,40,42,44,46,47,49,55–57,60^. The incidence of contralateral ischemic lesions remains significantly lower after the CEA procedures (1.4%, 9/649) compared to CAS technique (12.3%, 75/612) (OR: 0.16, 95% CI 0.08-0.32, p<0.001). The statistical heterogeneity was low (I^2^= 00.00%) **(Supplemental Figure 8)**.

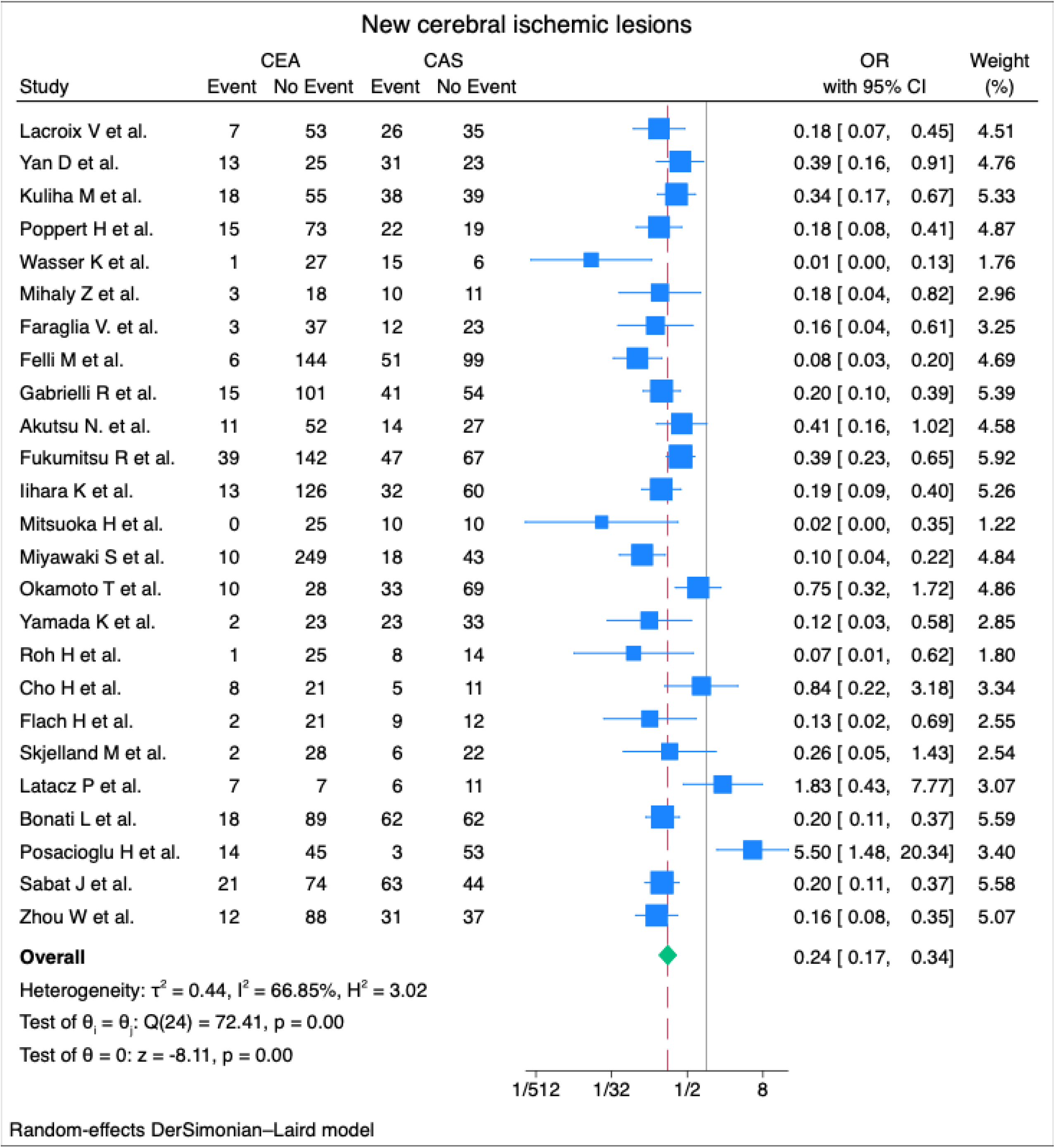

##### Stroke

The overall stroke rate was available in 17 studies including 28 stroke events after CEA (1.6%, 23/1429) and versus 53 events in the CAS group (4.7%, 53/1124)^27,43–47,49,51,52,54–59,61,62^. The stroke rate was significantly lower in CEA group compared to CAS technique (OR: 0.38, 95% CI: 0.23-0.63, p<0.001). The statistical heterogeneity was low (I^2^ = 00.00%) **(Figure 5)**.

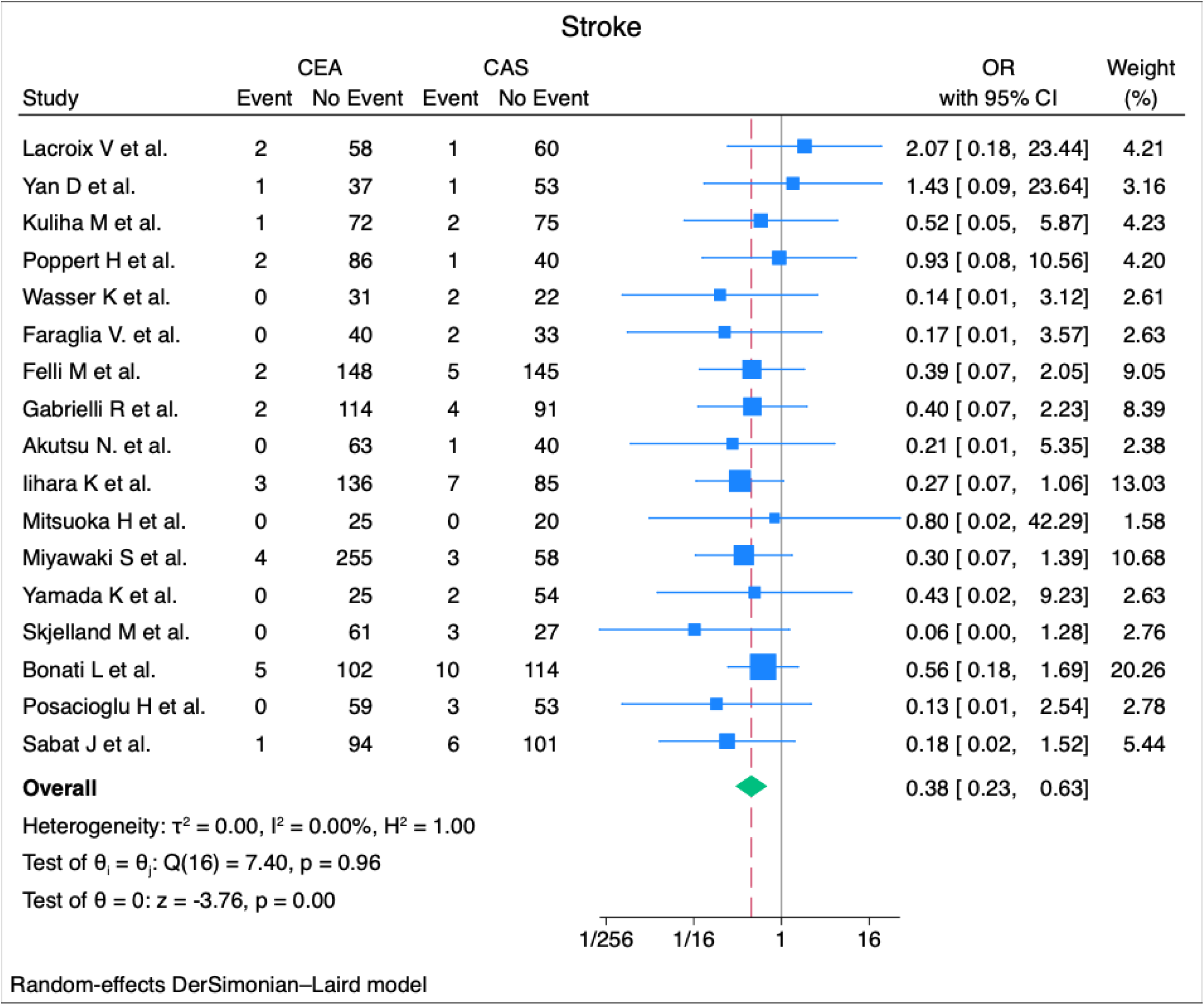

### Subgroup analysis

#### Study design

The incidence of new cerebral ischemic lesions was significantly lower after CEA comparing to CAS both in RCTs or PSM (CEA 21.4%, 46/215 vs CAS 48.5%, 116/239, OR: 0.34, 95% CI:0.16-0.75, p=0.01) and non-randomized studies (CEA 12.7 %, 205/1612 vs CAS 39.6%, 500/1261, OR: 0.23 95% CI:0.15-0.33, p<0.001). The statistical heterogeneity remained substantial in both RCTs or PSM (I^2^= 62.98%) and non-randomized studies (I^2^= 68.55%) **(Supplemental Figure 9)**. There was no statistically significant difference between these two subgroups (p=0.35).

### Embolic protection device

Three studies reported on data regarding the CAS technique without the use of protection device^27,44,51^. The CEA group (15.2%, 34/223) presented significantly lower rates of new ischemic cerebral lesions compared to CAS (44.5%, 57/128) (OR: 0.16, 95% CI: 0.05-0.54, p<0.001). The heterogeneity was substantial (I^2^= 74.74%) **(Supplemental Figure 10)**. Data concerning the CAS technique with the use of embolic protection device were extracted from 19 studies^27,33,40,42,43,45–47,49,51–56,58,59,62,63^. The CEA group (11.9%, 167/1403) presented significantly lower rates of new ischemic cerebral lesions compared to CAS (40.7%, 454/1116) (OR: 0.21, 95% CI: 0.14-0.34, p<0.001). The statistical heterogeneity remained substantial (I^2^= 73.64%) **(Supplemental Figure 10)**. There was no statistically significant difference between these two subgroups (p=0.65).

### Meta-regression analysis

The presence of pre-operative symptoms was expressed as the difference in rates of occurrence in the CEA versus the CAS group, and data were extracted from 19 studies^27,40,44–47,49,51–59,61–63^. Meta-regression analysis revealed that the presence of pre-operative symptoms had no statistically significant influence on the incidence of new cerebral ischemic lesions (*p*=0.241) **(Supplemental Figure 11)**. The data regarding hypertension were available in 15 studies^27,40,44–46,48,51,52,54–57,59,62,63^, and no statistically significant influence was found in the presence of hypertension and the rate of newly detected lesions (p=0.134) **(Supplemental Figure 12)**.The occurrence rates of diabetes and smoking were extracted from 15 studies^27,40,44–46,48,51,52,54–57,59,62,63^ and 13 studies^27,40,44–46,51,52,56,57,59,61–63^, respectively, and no statistically significant influence was detected between smoking (p=0.913) and diabetes (p=0.881) in the incidence of new cerebral ischemic lesion **(Supplemental Figure 13 and Supplemental Figure 14).**

## Discussion

Our study included a total of 25 studies reporting on new cerebral ischemic lesions after 1827 CEA procedures and 1500 CAS interventions. The results of this meta-analysis suggest that the incidence of new cerebral ischemic lesions was significantly lower in the CEA group compared to CAS regardless the time of MRI assessment. To address the heterogeneity between studies, we performed subgroup analyses based on the type of the study, RCTs or PSM and non-randomized, and the incidence of new lesions remained significantly higher after CAS. Moreover, the use of embolic protection device within the CAS procedure did not change the direction of the results, indicating significantly lower rates of new ischemic lesions after CEA.

The results of this study are in accordance with the previous meta-analysis of Gargiulo G et al., which is reporting on total 2014 procedures, and suggest slightly higher rates of incidence of new lesions in cerebral MRI (13.7% vs 12.2% in CEA and 41.1% vs 40.3% in CAS)^15^.

Furthermore, the meta-analysis of Schnaudigel S et al. included only studies that performed the MRI within 72 hours after the intervention^14^. This methodological approach does not allow to address the entirety of the available in the literature data concerning the new cerebral ischemic lesions. This might explain the low comparable rates of stroke events between CEA and CAS. They also included studies without direct comparison between the two techniques. The statistical analysis of these data seems to be inaccurate due to inadequate control of heterogeneity among the patients involved. Moreover, the previous meta-analysis of Traenka C et al., including 19 comparative studies, suggests a lower incidence of new cerebral ischemic lesions after CEA (10.6%, 118/1107) and CAS (40.4%, 401/993)^13^. The use of the fixed effect model for the calculation of the pooled effect may account for the slightly lower incidence observed in these findings compared to the results from our most updated meta-analysis with a total of 25 studies and incidence rates 13.7% (251/1827) and 41.1 (616/1500) after CEA and CAS, respectively.

Our study is the first to address the time variable, reporting on the interval between the operation and the subsequent MRI performance. Interestingly, the incidence of the new lesions within the first day following the revascularization (14.3% in CEA vs 41.3% in CAS) remains almost unchanged compared to 72 hours (13.6% in CEA vs 40.4% in CAS) and 1 week post-operatively (13.7% in CEA vs 41.1 in CAS). Our metanalysis is also the first to report on the contralateral cerebral ischemic lesions and the subgroup analysis concluded in lower rates after CEA compared to CAS. Embolic events could have occurred at any point during the revascularization, including angiography preceding CAS^64,65^. The dislodgment of thrombotic material or atherosclerotic debris during the stenting procedure, with the manipulation of the catheters, the sheaths and the guidewires in the supra-aortic arch section, seems to give rise to single or multiple emboli, leading potentially to the presence of lesions in MRI, even in the contralateral arterial distribution^66^. This may be an insight that a percentage of microemboli during CAS are indeed deriving from arch manipulations, and thus their distribution is bilateral; further research will shed some light upon this fact.

A substudy of the International Carotis Stenting Study (ICSS)^67^, reporting on the MRI findings, suggests an incidence 22.5% of stroke or transient ischemic attack in patients with new cerebral lesions in MRI versus 8.8% in patients without such lesions, following CAS^26^. The respective rates following the CEA were 11% versus 9%, during 5 years of follow up. The above results suggest that the silent micro-embolism detected on early MRI after CAS may indicate a higher risk of cerebrovascular events^26^. However, the ICSS included only patients with symptomatic carotid stenosis^67^. The impact of periprocedural MRI lesions in carotid revascularization in pre-operative asymptomatic patients on the risk of cerebrovascular events remains still unknown^5^. Likewise, the study of Traenka C et al., using a complex and controversial statistical methodology^68^, indicates a strong correlation between new cerebral ischemic lesions in the MRI and the probability of post procedural stroke event, in the combined CEA and CAS procedures^13^. The lack of adequate data within the studies included in our analysis prevents us from investigating the impact of the new cerebral ischemic lesions on the post-procedural clinical events and cognitive impairment.

To address the heterogeneity in CAS procedure concerning the use of an embolic protection device, we performed a subgroup analysis. The results of this analysis suggest slightly lower rates of new ischemic cerebral lesions following the implementation of an embolic protection device, with rates of 40.7% compared to 44.5% without any protection device. It is important to highlight that microeboli still can overcome embolic protection devices. The meta-analysis of Traenka C et al reports lower rates of occurrence of ischemic lesions, following the CAS technique, 36.8% with protection versus 37.1% without protection device^13^. In any case, the incidence of silent microembolism remains statistically significantly lower irrespective of whether an embolic protection device was used or not. Our findings appear to align also with the reported incidence of 37% of the CARENET Trial (Carotid Embolic Protection Using MicroNet)^69^. The aforementioned result may indicate that the dislodgement of embolic particles is not adequately prevented by the use embolic protection device. Interestingly, recent prospective multicentre study concerning the CGuard MicroNet-covered Embolic Prevention System reported 19.6% new ischemic microembolisation in MRI performed within 72 hours post intervention^70^. The limited available data concerning the stent type in the CAS group prevents us from proceeding to further analysis regarding the new cerebral ischemic lesions in these subgroups.

The preoperative cardiovascular risk along with the characteristics of the atherosclerotic plaque and the severity of stenosis are both unknown and imbalanced between the two groups. To address this issue, a subgroup analysis of RCTs and non-randomized was performed, and the findings are consistent with the overall result, indicating a lower incidence of new cerebral lesions following CEA. Furthermore, the meta-regression analysis concerning the pre-operative symptomatic status, hypertension, smoking and diabetes did not show a significant impact of these moderators in the difference of the incidence between the two techniques. These results are in accordance with the findings of a non-randomized study addressing symptomatic carotid stenosis treated with CAS, which demonstrated no correlation between these variables and the presence of cerebral ischemic lesions^71^. One retrospective study indicated the diabetes mellitus as a predictor of perioperative stroke or death among patients undergoing CEA, whereas it did not exhibit the same predictive significance among patients undergoing CAS^72^.

The limitations of the research should be acknowledged. First, there were the inherent limitations of a study-level meta-analysis as we did not have access to patient-level covariates that could confound our findings. Most of the studies do not mention many important periprocedural data that could affect the final outcomes: the rate of pre-existing contralateral carotid stenosis, use of eversion technique, shunting, the clamping duration and the operation time, and the hemodynamic parameters. Moreover, it is noteworthy to mention that variations in magnetic field strengths may have contributed to differences in the sensitivity of detecting small ischemic brain lesions. In addition, there is inadequate information regarding the volume and the number of the lesions. Furthermore, the 22 of the 25 included studies are not randomized. The lack of randomization, allocation concealment, and blinding increases the chances of confounding and selection bias. Considering that CAS is usually performed on high-risk patients, the reported results may not be generalizable to other patients with intermediate surgical risk. Likewise, all the procedures in the included studies were performed in different centers by different operators. This fact, along with differences in the follow-up schemes across the studies, might have affected the generalizability of our results.

## Conclusions

This meta-analysis indicates that CEA is associated with a significantly lower incidence of new cerebral ischemic lesions in early post-operative MRI, compared to CAS, regardless of whether they are ipsilateral or contralateral to the respective carotid artery. Embolic protection devices did not alter the outcome. The clinical importance of the incidence of silent cerebral microemboli as predictive factors for post-operative clinical outcomes along with emerging stenting techniques and types, remains to be defined.

## Data Availability

We affirm that all data referred to in the manuscript are readily available upon request

## Acknowledgment

None

## References

1. Antonopoulos CN, Kakisis JD, Sfyroeras GS, et al. The impact of carotid artery stenting on cognitive function in patients with extracranial carotid artery stenosis. Ann Vasc Surg. 2015;29(3):457–469. doi:10.1016/j.avsg.2014.10.024

2. Kim JJ, Schwartz S, Wen J, et al. Comparison of Neurocognitive Outcomes after Carotid Endarterectomy and Carotid Artery Stenting. Am Surg. 2015;81(10):1010–1014.

3. Lal BK, Younes M, Cruz G, Kapadia I, Jamil Z, Pappas PJ. Cognitive changes after surgery vs stenting for carotid artery stenosis. J Vasc Surg. 2011;54(3):691–698. doi:10.1016/j.jvs.2011.03.253

4. Sabat J, Bock D, Hsu CH, et al. Risk factors associated with microembolization after carotid intervention. J Vasc Surg. 2020;71(5):1572–1578. doi:10.1016/j.jvs.2019.06.202

5. Naylor R, Rantner B, Ancetti S, et al. Editor’s Choice - European Society for Vascular Surgery (ESVS) 2023 Clinical Practice Guidelines on the Management of Atherosclerotic Carotid and Vertebral Artery Disease. Eur J Vasc Endovasc Surg. 2023;65(1):7–111. doi:10.1016/j.ejvs.2022.04.011

6. Montorsi P, Caputi L, Galli S, et al. Microembolization during carotid artery stenting in patients with high-risk, lipid-rich plaque. A randomized trial of proximal versus distal cerebral protection. J Am Coll Cardiol. 2011;58(16):1656–1663. doi:10.1016/j.jacc.2011.07.015

7. Cano MN, Kambara AM, de Cano SJF, et al. Randomized comparison of distal and proximal cerebral protection during carotid artery stenting. JACC Cardiovasc Interv. 2013;6(11):1203–1209. doi:10.1016/j.jcin.2013.07.006

8. Texakalidis P, Letsos A, Kokkinidis DG, et al. Proximal embolic protection versus distal filter protection versus combined protection in carotid artery stenting: A systematic review and meta-analysis. Cardiovasc Revasc Med. 2018;19(5 Pt A):545-552. doi:10.1016/j.carrev.2017.12.010

9. Bowden D, Hayes N, London N, Bell P, Naylor AR, Hayes P. Carotid endarterectomy performed in the morning is associated with increased cerebral microembolization. J Vasc Surg. 2009;50(1):48–53. doi:10.1016/j.jvs.2009.01.011

10. Hitchner E, Baughman BD, Soman S, Long B, Rosen A, Zhou W. Microembolization is associated with transient cognitive decline in patients undergoing carotid interventions. J Vasc Surg. 2016;64(6):1719–1725. doi:10.1016/j.jvs.2016.06.104

11. Varetto G, Gibello L, Faletti R, et al. Contrast-enhanced ultrasound to predict the risk of microembolization during carotid artery stenting. Radiol Med. 2015;120(11):1050–1055. doi:10.1007/s11547-015-0530-4

12. Hammer FD, Lacroix V, Duprez T, et al. Cerebral microembolization after protected carotid artery stenting in surgical high-risk patients: results of a 2-year prospective study. J Vasc Surg. 2005;42(5):847–853; discussion 853. doi:10.1016/j.jvs.2005.05.065

13. Traenka C, Engelter ST, Brown MM, Dobson J, Frost C, Bonati LH. Silent brain infarcts on diffusion-weighted imaging after carotid revascularisation: A surrogate outcome measure for procedural stroke? A systematic review and meta-analysis. Eur Stroke J. 2019;4(2):127–143. doi:10.1177/2396987318824491

14. Schnaudigel S, Gröschel K, Pilgram SM, Kastrup A. New brain lesions after carotid stenting versus carotid endarterectomy: a systematic review of the literature. Stroke. 2008;39(6):1911–1919. doi:10.1161/STROKEAHA.107.500603

15. Gargiulo G, Sannino A, Stabile E, Perrino C, Trimarco B, Esposito G. New cerebral lesions at magnetic resonance imaging after carotid artery stenting versus endarterectomy: an updated meta-analysis. PLoS One. 2015;10(5):e0129209. doi:10.1371/journal.pone.0129209

16. Page MJ, McKenzie JE, Bossuyt PM, et al. The PRISMA 2020 statement: an updated guideline for reporting systematic reviews. BMJ. 2021;372:n71. doi:10.1136/bmj.n71

17. Wohlin C. Guidelines for Snowballing in Systematic Literature Studies and a Replication in Software Engineering. In: Proceedings of the 18th International Conference on Evaluation and Assessment in Software Engineering. EASE ‘14. Association for Computing Machinery; 2014. doi:10.1145/2601248.2601268

18. Covidence systematic review software, Veritas Health Innovation, Melbourne, Australia. Available at www.covidence.org.

19. Higgins JPT, Altman DG, Gøtzsche PC, et al. The Cochrane Collaboration’s tool for assessing risk of bias in randomised trials. BMJ. 2011;343:d5928. doi:10.1136/bmj.d5928

20. Sterne JA, Hernán MA, Reeves BC, et al. ROBINS-I: a tool for assessing risk of bias in non-randomised studies of interventions. BMJ. 2016;355:i4919. doi:10.1136/bmj.i4919

21. Hozo SP, Djulbegovic B, Hozo I. Estimating the mean and variance from the median, range, and the size of a sample. BMC Med Res Methodol. 2005;5:13. doi:10.1186/1471-2288-5-13

22. Wan X, Wang W, Liu J, Tong T. Estimating the sample mean and standard deviation from the sample size, median, range and/or interquartile range. BMC Med Res Methodol. 2014;14:135. doi:10.1186/1471-2288-14-135

23. Sweeting MJ, Sutton AJ, Lambert PC. What to add to nothing? Use and avoidance of continuity corrections in meta-analysis of sparse data. Stat Med. 2004;23(9):1351–1375. doi:10.1002/sim.1761

24. DerSimonian R, Laird N. Meta-analysis in clinical trials. Control Clin Trials. 1986;7(3):177–188. doi:10.1016/0197-2456(86)90046-2

25. Gensicke H, Zumbrunn T, Jongen LM, et al. Characteristics of ischemic brain lesions after stenting or endarterectomy for symptomatic carotid artery stenosis: Results from the international carotid stenting study-magnetic resonance imaging substudy. Stroke. 2013;44(1):80–86. doi:10.1161/STROKEAHA.112.673152

26. Gensicke H, Van Der Worp HB, Nederkoorn PJ, et al. Ischemic brain lesions after carotid artery stenting increase future cerebrovascular risk. J Am Coll Cardiol. 2015;65(6):521–529. doi:10.1016/j.jacc.2014.11.038

27. Bonati LH, Jongen LM, Haller S, et al. New ischaemic brain lesions on MRI after stenting or endarterectomy for symptomatic carotid stenosis: a substudy of the International Carotid Stenting Study (ICSS). Lancet Neurol. 2010;9(4):353–362. doi:10.1016/S1474-4422(10)70057-0

28. Müller MD, Ahlhelm FJ, Von Hessling A, et al. Vascular Anatomy Predicts the Risk of Cerebral Ischemia in Patients Randomized to Carotid Stenting Versus Endarterectomy. Stroke. 2017;48(5):1285–1292. doi:10.1161/STROKEAHA.116.014612

29. Altinbas A, Van Zandvoort MJE, Van Den Berg E, et al. Cognition after carotid endarterectomy or stenting: A randomized comparison. Neurology. 2011;77(11):1084–1090. doi:10.1212/WNL.0b013e31822e55b9

30. Rostamzadeh A, Zumbrunn T, Jongen LM, et al. Predictors of acute and persisting ischemic brain lesions in patients randomized to carotid stenting or endarterectomy. Stroke. 2014;45(2):591–594. doi:10.1161/STROKEAHA.113.003605

31. Burow A, Lyrer PA, Nederkoorn PJ, et al. Echographic risk index and cerebral ischemic brain lesions in patients randomized to stenting versus endarterectomy for symptomatic carotid artery stenosis. Ultraschall in der Medizin. 2014;35(3):267–272. doi:10.1055/s-0033-1355751

32. Donners SJA, Rots ML, Toorop RJ, Van Der Lugt A, Bonati LH, De Borst GJ. Long-Term Stroke Risk in Patients with New Ischemic Brain Lesions on MRI after Carotid Revascularization. Stroke. 2023;54(10):2562–2568. doi:10.1161/STROKEAHA.123.043336

33. Zhou W, Dinishak D, Lane B, Hernandez-Boussard T, Bech F, Rosen A. Long-term radiographic outcomes of microemboli following carotid interventions. J Vasc Surg. 2009;50(6):1314–1319. doi:10.1016/j.jvs.2009.07.105

34. Zhou W, Baughman BD, Soman S, et al. Volume of subclinical embolic infarct correlates to long-term cognitive changes after carotid revascularization. J Vasc Surg. 2017;65(3):686–694. doi:10.1016/j.jvs.2016.09.057

35. Tedesco MM, Lee JT, Dalman RL, et al. Postprocedural microembolic events following carotid surgery and carotid angioplasty and stenting. J Vasc Surg. 2007;46(2):244–250. doi:10.1016/j.jvs.2007.04.049

36. Hitchner E, Baughman BD, Soman S, Long B, Rosen A, Zhou W. Microembolization is associated with transient cognitive decline in patients undergoing carotid interventions. J Vasc Surg. 2016;64(6):1719–1725. doi:10.1016/j.jvs.2016.06.104

37. Zhou W, Hitchner E, Gillis K, et al. Prospective neurocognitive evaluation of patients undergoing carotid interventions. J Vasc Surg. 2012;56(6):1571–1578. doi:10.1016/j.jvs.2012.05.092

38. Kraemer C, Nisson P, Wheeler G, et al. Patient risk factors associated with embolic stroke volumes after revascularization. J Vasc Surg. 2020;72(6):2061–2068. doi:10.1016/j.jvs.2020.02.040

39. Tedesco MM, Coogan SM, Dalman RL, et al. Risk factors for developing postprocedural microemboli following carotid interventions. Journal of Endovascular Therapy. 2007;14(4):561–567. doi:10.1583/1545-1550(2007)14[561:RFFDPM]2.0.CO;2

40. Mihály Z, Booth S, Nguyen DT, et al. A Propensity-Matched Comparison of Ischemic Brain Lesions on Postprocedural MRI in Endovascular versus Open Carotid Artery Reconstruction. J Cardiovasc Dev Dis. 2023;10(6). doi:10.3390/jcdd10060257

41. García-Sánchez S, Millán-Torné M, Capellades-Font J, Muchart J, Callejas-P JM, Vila-Moriente N. [Ischemic brain lesions following carotid revascularisation procedures: a comparative study using diffusion-weighted magnetic resonance imaging]. Rev Neurol. 2004;38(11):1013–1017.

42. Latacz P, Simka M, Bryll A, Piwowarczyk M, Gajos G, Popiela T. Cerebral ischemic lesions on diffusionweighted magnetic resonance imaging after carotid eversion endarterectomy vs carotid stenting with a proximal protection device: Results of a randomized prospective trial. Pol Arch Intern Med. 2019;129(7-8):562–566. doi:10.20452/pamw.14825

43. Felli MMG, Alunno A, Castiglione A, et al. CEA versus CAS: Short-term and mid-term results. International Angiology. 2012;31(5):420–426.

44. Poppert H, Wolf O, Resch M, et al. Differences in number, size and location of intracranial microembolic lesions after surgical versus endovascular treatment without protection device of carotid artery stenosis. J Neurol. 2004;251(10):1198–1203. doi:10.1007/s00415-004-0502-4

45. Sabat J, Bock D, Hsu CH, et al. Risk factors associated with microembolization after carotid intervention. J Vasc Surg. 2020;71(5):1572–1578. doi:10.1016/j.jvs.2019.06.202

46. Gabrielli R, Siani A, Smedile G, Rizzo AR, Accrocca F, Bartoli S. Carotid Artery Stenting versus Carotid Endarterectomy in Terms of Neuroprotection DW-MRI Detected and Neuropsychological Assessment Impairment. Ann Vasc Surg. 2024;98(June 2023):68–74. doi:10.1016/j.avsg.2023.05.046

47. Lacroix V, Hammer F, Astarci P, et al. Ischemic Cerebral Lesions after Carotid Surgery and Carotid Stenting. European Journal of Vascular and Endovascular Surgery. 2007;33(4):430–435. doi:10.1016/j.ejvs.2006.11.012

48. Fukumitsu R, Yoshida K, Kurosaki Y, et al. Short-Term Results of Carotid Endarterectomy and Stenting After the Introduction of Carotid Magnetic Resonance Imaging: A Single-Institution Retrospective Study. World Neurosurg. 2017;101:308–314. doi:10.1016/j.wneu.2017.02.032

49. Akutsu N, Hosoda K, Fujita A, Kohmura E. A preliminary prediction model with MR plaque imaging to estimate risk for new ischemic brain lesions on diffusion-weighted imaging after endarterectomy or stenting in patients with carotid stenosis. American Journal of Neuroradiology. 2012;33(8):1557–1564. doi:10.3174/ajnr.A3002

50. Cho AH, Cho YP, Lee DH, et al. Reperfusion injury on magnetic resonance imaging after carotid revascularization. Stroke. 2014;45(2):602–604. doi:10.1161/STROKEAHA.113.003792

51. Wasser K, Pilgram-Pastor SM, Schnaudigel S, et al. New brain lesions after carotid revascularization are not associated with cognitive performance. J Vasc Surg. 2011;53(1):61–70. doi:10.1016/j.jvs.2010.07.061

52. Yan D, Tang X, Shi Z, et al. Perioperative and Follow-up Results of Carotid Artery Stenting and Carotid Endarterectomy in Patients with Carotid Near-Occlusion. Ann Vasc Surg. 2019;59:21–27. doi:10.1016/j.avsg.2019.01.019

53. Roh HG, Byun HS, Ryoo JW, et al. Prospective analysis of cerebral infarction after carotid endarterectomy and carotid artery stent placement by using diffusion-weighted imaging. American Journal of Neuroradiology. 2005;26(2):376–384.

54. Miyawaki S, Maeda K. Surgical treatment for cervical carotid artery stenosis in the elderly: Importance of perioperative management of ischemic cardiac complications. Neurol Med Chir (Tokyo). 2014;54(2):120–125. doi:10.2176/nmc.oa2012-0436

55. Yamada K, Yoshimura S, Kawasaki M, et al. Embolic complications after carotid artery stenting or carotid endarterectomy are associated with tissue characteristics of carotid plaques evaluated by magnetic resonance imaging. Atherosclerosis. 2011;215(2):399–404. doi:10.1016/j.atherosclerosis.2011.01.002

56. Posacioglu H, Engin C, Cinar C, et al. Carotid endarterectomy versus carotid artery stenting: Findings in regard to neuroclinical outcomes and diffusion-weighted imaging. Tex Heart Inst J. 2008;35(4):395–401.

57. Kuliha M, Roubec M, Procházka V, et al. Randomized clinical trial comparing neurological outcomes after carotid endarterectomy or stenting. British Journal of Surgery. 2015;102(3):194–201. doi:10.1002/bjs.9677

58. Mitsuoka H, Shintani T, Furuya H, Nakao Y, Higashi S. Ultrasonographic Character of Carotid Plaque and Postprocedural Brain Embolisms in Carotid Artery Stenting and Carotid Endarterectomy. Ann Vasc Dis. 2011;4(2):106–109. doi:10.3400/avd.oa.11.00007

59. Iihara K, Murao K, Sakai N, Yamada N, Nagata I, Miyamoto S. Outcome of carotid endarterectomy and stent insertion based on grading of carotid endarterectomy risk: A 7-year prospective study. J Neurosurg. 2006;105(4):546–554. doi:10.3171/jns.2006.105.4.546

60. Flach HZ, Ouhlous M, Hendriks JM, et al. Cerebral ischemia after carotid intervention. Journal of Endovascular Therapy. 2004;11(3):251–257. doi:10.1583/03-1128.1

61. Skjelland M, Krohg-Sørensen K, Tennøe B, Bakke SJ, Brucher R, Russell D. Cerebral microemboli and brain injury during carotid artery endarterectomy and stenting. Stroke. 2009;40(1):230–234. doi:10.1161/STROKEAHA.107.513341

62. Faraglia V, Palombo G, Stella N, et al. Cerebral embolization in patients undergoing protected carotid-artery stenting and carotid surgery. J Cardiovasc Surg (Torino). 2007;48(6):683–688.

63. Okamoto T, Inoue Y, Oi Y, et al. Strategy of carotid artery stenting as first-line treatment and carotid endarterectomy for carotid artery stenosis: A single-center experience. Surg Neurol Int. 2022;13:513. doi:10.25259/SNI_820_2022

64. Bendszus M, Koltzenburg M, Burger R, Warmuth-Metz M, Hofmann E, Solymosi L. Silent embolism in diagnostic cerebral angiography and neurointerventional procedures: a prospective study. Lancet. 1999;354(9190):1594–1597. doi:10.1016/S0140-6736(99)07083-X

65. Gerraty RP, Bowser DN, Infeld B, Mitchell PJ, Davis SM. Microemboli during carotid angiography. Association with stroke risk factors or subsequent magnetic resonance imaging changes? Stroke. 1996;27(9):1543–1547. doi:10.1161/01.str.27.9.1543

66. Orlandi G, Fanucchi S, Fioretti C, et al. Characteristics of Cerebral Microembolism During Carotid Stenting and Angioplasty Alone. Arch Neurol. 2001;58(9):1410–1413. doi:10.1001/archneur.58.9.1410

67. Bonati LH, Dobson J, Featherstone RL, et al. Long-term outcomes after stenting versus endarterectomy for treatment of symptomatic carotid stenosis: The International Carotid Stenting Study (ICSS) randomised trial. The Lancet. 2015;385(9967):529–538. doi:10.1016/S0140-6736(14)61184-3

68. Fleming TR, Powers JH. Biomarkers and surrogate endpoints in clinical trials. Stat Med. 2012;31(25):2973–2984. doi:10.1002/sim.5403

69. Schofer J, Musiałek P, Bijuklic K, et al. A Prospective, Multicenter Study of a Novel Mesh-Covered Carotid Stent: The CGuard CARENET Trial (Carotid Embolic Protection Using MicroNet). JACC Cardiovasc Interv. 2015;8(9):1229–1234. doi:10.1016/j.jcin.2015.04.016

70. Speziale F, Capoccia L, Sirignano P, et al. Thirty-day results from prospective multi-specialty evaluation of carotid artery stenting using the CGuard MicroNet-covered Embolic Prevention System in real-world multicentre clinical practice: the IRON-Guard study. EuroIntervention. 2018;13(14):1714–1720. doi:10.4244/EIJ-D-17-00008

71. Russjan A, Goebell E, Havemeister S, et al. Predictors of periprocedural brain lesions associated with carotid stenting. Cerebrovasc Dis. 2012;33(1):30–36. doi:10.1159/000332088

72. Parlani G, De Rango P, Cieri E, et al. Diabetes is not a predictor of outcome for carotid revascularization with stenting as it may be for carotid endarterectomy. J Vasc Surg. 2012;55(1):79. doi:10.1016/j.jvs.2011.07.080

